# Distinguishing benign from pathogenic duplications involving *GPR101* and *VGLL1*-adjacent enhancers in the clinical setting with the bioinformatic tool POSTRE

**DOI:** 10.1101/2025.06.27.25329768

**Authors:** Giampaolo Trivellin, Víctor Sánchez-Gaya, Alexia Grasso, Magdalena Pasińska, Constantine A. Stratakis, Di Milnes, Edwin P. Kirk, Albert Beckers, Andrea G. Lania, Patrick Pétrossians, Alvaro Rada-Iglesias, Martin Franke, Adrian F. Daly

**Affiliations:** Department of Biomedical Sciences, Humanitas University, via Rita Levi Montalcini 4, 20072 Pieve Emanuele, Milan, Italy; IRCCS Humanitas Research Hospital, Translational Endocrinology and Metabolism Lab, via Manzoni 56, 20089 Rozzano, Milan, Italy; Institute of Biomedicine and Biotechnology of Cantabria (IBBTEC), CSIC/Universidad de Cantabria, Albert Einstein 22, 39011 Santander, Spain; Department of Clinical Genetics, Faculty of Medicine, Collegium Medicum in Bydgoszcz, Nicolaus Copernicus University, Bydgoszcz, Poland; Human Genetics & Precision Medicine, Institute of Molecular Biology and Biotechnology (IMBB), Foundation for Research and Technology Hellas (FORTH), Heraklion, Greece; Genetic Health Queensland, Royal Brisbane Women’s Hospital, Brisbane, QLD Australia; NSW Health Pathology East Genomics, Randwick, NSW, Australia; School of Clinical Medicine, University of New South Wales, Randwick NSW, Australia; Department of Endocrinology, Centre Hospitalier Universitaire de Liège, University of Liège, 4000 Liège, Belgium; Andalusian Center for Developmental Biology (CABD), Junta de Andaluciıa– Universidad Pablo de Olavide (UPO) – Consejo Superior de Investigaciones Científicas (CSIC), Seville, Spain

**Keywords:** topologically associating domains (TADs), chromatin architecture, prenatal diagnostics, copy number variants (CNVs), X-linked acrogigantism (X-LAG)

## Abstract

**Background:** Structural variants (SVs) that disrupt topologically associating domains (TADs) can cause disease by rewiring enhancer-promoter interactions. Duplications involving *GPR101* are known to cause X-linked acrogigantism (X-LAG) by enabling aberrant expression of *GPR101* through hijacking of enhancers at *VGLL1*. However, not all *GPR101*-containing duplications are pathogenic, presenting a diagnostic challenge, especially in the prenatal setting.

**Methods:** We evaluated POSTRE, a tool designed to predict the regulatory impact of SVs, to distinguish pathogenic from benign *GPR101* duplications. We analyzed six non-pathogenic duplications, and 27 known X-LAG associated pathogenic duplications. Tissue-specific enhancer maps built using H3K27ac ChIP-seq and ATAC-seq data as well as gene expression data derived from human anterior pituitary samples were integrated into POSTRE to enable predictions in a X-LAG relevant tissue context.

**Results:** POSTRE correctly classified all 33 duplications as benign or pathogenic. In addition, one X-LAG case with mild clinical features (e.g., severe GH hypersecretion in the absence of pituitary tumorigenesis) was found to include only 2/5 *VGLL1* enhancers (also predicted to be the weakest enhancers), whereas all 26 typical X-LAG cases had 24 enhancers duplicated. This suggests that milder enhancer hijacking at *VGLL1* could explain the different clinical features of X-LAG in this individual.

**Conclusions:** These findings support the utility of POSTRE to support diagnostic pipelines when interpreting SVs affecting chromatin architecture in pituitary disease. By accurately modelling enhancer adoption in a cell type-specific context, POSTRE could help to reduce uncertainty in genetic counselling and offers a rapid alternative to performing chromatin conformation capture experiments.

## Introduction

The spatial organization of the genome plays a fundamental role in gene regulation. At the sub-mega-base scale, chromatin architecture is partitioned into topologically associating domains (TADs)^1^. These are self-interacting regions that constrain enhancer-promoter communications and insulate genes from regulatory elements outside their TAD^1–3^. Disruption of TADs by structural variants (SVs), such as duplications, deletions, inversions, or translocations can result in aberrant gene expression by altering the regulatory landscape, a phenomenon increasingly recognized as a common mechanism in human disease^3–7^. These so-called “TADopathies”^8^ have been implicated in a growing number of disorders, highlighting the importance of chromatin architecture in maintaining gene expression fidelity^9^.

One TADopathy is X-linked acrogigantism (X-LAG), a very rare condition characterized by early pediatric growth hormone (GH) excess due to pituitary tumors and/or hyperplasia^10^. The disease is caused by duplications at chromosome Xq26.3 involving the *GPR101* gene. We demonstrated recently that these duplications can disrupt the local *GPR101* TAD structure and lead to the formation of a pathological neo-TAD. The neo-TAD brings *GPR101* into contact with ectopic enhancers, resulting in its pathological overexpression^11,12^. A critical ectopic enhancer cluster implicated in this mechanism resides within the *VGLL1* gene locus and is active specifically in pituitary cells^12^. The action of this element on *GPR101* likely underlies the pathogenicity of duplications in X-LAG.

Despite the growing understanding of these regulatory mechanisms, distinguishing pathogenic from benign duplications involving *GPR101* remains a major diagnostic challenge. This is especially true in the context of prenatal chromosomal microarray analysis (CMA), where duplications may be detected incidentally, creating the potential for significant distress, due to the challenges inherent in fetal phenotyping. Without functional data, interpreting the clinical significance of these findings is difficult, often leading to uncertainty in genetic counseling and clinical decision-making^13^. We recently showed that 4C-seq and HiC can map chromatin interactions and assess TAD integrity in these, thereby playing a diagnostic role^14^. Notwithstanding their clinical utility, we are aware that these techniques are highly technically specialized, labor-intensive, and not readily available in most clinical laboratories.

In this context, there is a pressing need for computational approaches capable of predicting the regulatory impact of structural variants using only the SV genomic coordinates and accessible omics data (e.g., HiC, ChIP-Seq, ATAC-seq, RNA-seq). One such approach is POSTRE (Prediction Of STRuctural variant Effects), a recently developed tool designed to model SV long-range pathomechanisms, including enhancer adoption and neo-TAD formation, based on tissue-specific genomic data. By integrating TAD maps with chromatin accessibility, histone modifications, and gene expression data, POSTRE enables the prediction of regulatory disruptions caused by SVs in phenotypically-relevant cellular contexts^15^. As originally implemented, POSTRE was only applicable to a limited set of congenital defects (i.e. neurodevelopmental, craniofacial, limb, heart) for which genomic data in disease relevant human tissues was analyzed and integrated. In the present study, we developed an expanded version of POSTRE that is enriched with anterior pituitary-specific data, thus making it compatible with X-LAG and, potentially, with other disorders involving the pituitary gland. We then assessed whether POSTRE could accurately discriminate pathogenic from benign duplications at the *GPR101* locus and thereby serve as a tool to inform the clinical interpretation of TAD-disrupting SVs.

## Materials and Methods

### Study population

The study population consisted of 33 individuals, of whom 30 had previously reported pathogenic *GPR101* duplications associated with X-LAG^10,12,16–20^ or non-pathogenic duplications^14^. Three newly identified individuals were also included, they harbored microduplications at the *GPR101* locus on chromosome Xq26.3 that were discovered incidentally during prenatal or pediatric genetic testing at sites in three different countries. None of the three individuals exhibited signs of pituitary hyperplasia/tumor, gigantism, or other endocrine abnormalities consistent with X-LAG at the time of assessment. Individual F4A, a female, was identified via prenatal chromosomal microarray analysis (CMA) of DNA extracted from amniotic fluid that was performed for advanced maternal age. Individual F5A, a female, was identified after CMA on chorionic villus sampling for investigation of unrelated fetal ultrasound findings. Individual F6A was diagnosed postnatally as part of a clinical workup for developmental concerns without endocrine pathology. Some details of F6A were previously reported^21^.

### Copy number variant (CNV) analysis in new subjects

CMA was performed in F4A on embryonic DNA derived from amniotic fluid using the Agilent Technologies G3 ISCA V2 8×60K CGH microarray platform. This revealed a 366 kb duplication at Xq26.3, corresponding to genomic coordinates chrX:136,104,659-136,470,844 (hg19). The duplicated segment included the genes *RBMX* and *GPR101* but did not encompass the *VGLL1* gene or its intronic enhancer cluster, which we previously linked to the pathogenesis of X-LAG^12^. The duplication was inherited from a healthy parent, who had no history of growth or other disorders; she was born at term after a normal pregnancy and no evidence of growth disorders was noted.

DNA for F5A was extracted directly from blood and analyzed using the Illumina 850K CytoSNP v1.4 SNP microarray (50 kb mean effective resolution). F5A had a 237 kb duplication involving the region chrX:135,954,223-136,191,468 (hg19). This duplication also included *RBMX* and *GPR101*, but did not reach the *VGLL1* locus. F5A had no history of growth disorders and her hormone levels were normal.

Genomic DNAs from F6A and his healthy parent were analyzed using a clinical-grade 60K CGH array (Agilent ISCA CGH 60K, AMADID 031746), which provides genome-wide coverage with an estimated effective resolution of about 100 kb for gains and losses. The aCGH array analysis identified a 444.8 kb duplication spanning chrX:135,702,382-136,147,166 (hg19). aCGH analysis performed on F6A’s parent did not detect the duplication, indicating a *de novo* origin. Confirmation of the CNV was performed using droplet digital PCR (ddPCR), with probes targeting four genes located within the duplication (*CD40LG*, *ARHGEF6*, *RBMX*, and *GPR101*) and two control genes outside the duplicated segment (*HTATSF1* and *ZIC3*). DNA from a previously diagnosed X-LAG patient harboring a constitutional duplication served as a positive control for the ddPCR study. ddPCR showed that the duplication was mosaic, with an average copy number of 1.44 for five probes located within the duplicated region (Supplementary Figure 1). This indicated that approximately 44% of peripheral blood cells carried the duplication. The affected genes included *CD40LG*, *ARHGEF6*, *RBMX*, and *GPR101*, while flanking genes such as *HTATSF1* and *ZIC3* showed a diploid copy number.

### Genomic data processing (RNA-seq, ChIP-seq, ATAC-seq and HiC data) and incorporation into POSTRE

To enable accurate modeling of enhancer adoption and regulatory disruption in pituitary tissue, we generated a tissue-specific dataset (NormalPituitary) and supplemented this with publicly available transcriptomic data (NormalPituitary2) for cross-validation.

Normal adult anterior pituitary tissue samples (two males, one female) were obtained post-mortem through Cureline Inc, as previously reported^12^. These were used to generate RNA-seq, ATAC-seq, and H3K27ac ChIP-seq data. RNA sequencing was performed on total RNA extracted from these tissues (for further details refer to Franke et al.^12^). Prior to its incorporation into POSTRE, gene expression levels were converted to RPKMs and aggregated. In this regard, for each gene, the average expression across the three samples was taken as reference. In parallel, RNA-seq data normalized in the form of Transcripts Per Million (TPM)s, from normal whole pituitary glands were obtained from the Genotype-Tissue Expression (GTEx) Portal. In particular, the gene_tpm_v10_pituitary.gct.gz file was downloaded in December 2024 from the “Bulk tissue expression” section in the downloads page. Next, the median expression value for each gene across all available samples was taken as reference. The GTEx data formed the basis for the NormalPituitary2 condition in POSTRE, providing an additional independent gene expression match to the enhancer maps.

The TAD coordinates used by POSTRE, in NormalPituitary and NormalPituitary2, were derived from the brain prefrontal cortex boundary map provided from Schmitt et al.^22^ as previously done for other tissues (for more details please visit the POSTRE user guide). ChIP-seq profiling of the H3K27ac histone mark was performed on an additional normal adult anterior pituitary sample using the ChIP-IT High Sensitivity kit (Cat. No. V13-53040, Active Motif,) and a polyclonal anti-H3K27ac antibody (Cat. No. 39135, Active Motif). Library preparation was conducted using the Novogene NGS DNA Library Prep Set (Cat. No. PT004), and sequencing was performed on the Illumina NovaSeq X Plus platform with 150 bp paired-end reads.

ATAC-seq was performed on two anterior pituitary samples using about 50,000 cells per preparation. The ATAC-seq Kit (Cat. No. C01080006, Diagenode) was used according to the manufacturer’s protocol, including Illumina-compatible library generation and indexing (UDI Index Set I, Diagenode). Sequencing was carried out at Azenta Life Sciences using the Illumina NovaSeq platform (2×150 bp), yielding about 350 million paired-end reads (∼105 GB) per sample.

The resulting H3K27ac and ATAC-seq datasets were integrated to define tissue-specific active enhancers. In this regard, enhancers were defined as ATAC peaks (fold change>4 and q value <0.05) located more than 5 kb from any protein coding transcription start site (TSS) and within 500 bp of H3K27ac peaks (fold change >2 and p value <0.05). Finally, the enhancer maps were matched, in parallel, with the NormalPituitary and NormalPituitary2 gene expression datasets, giving rise to the two different conditions evaluated in POSTRE in relation with Endocrine abnormalities.

### In silico modeling with POSTRE

The pathogenic potential of multiple CNVs was assessed using the POSTRE software, a computational tool, available at https://github.com/vicsanga/Postre, designed to predict the regulatory impact of SVs that alter enhancer-promoter interactions^15^. To do so,

POSTRE utilizes gene expression and enhancer maps from disease relevant cell types and combines them with available TAD maps. Since the first version of POSTRE did not present genomic data relevant for the interpretation of pituitary related diseases, an expanded version has been created in this project, as described in the previous Method section. The expanded version, which includes the newly incorporated pituitary data, is currently available on request and will be directly available through POSTRE GitHub page upon peer-reviewed publication of this manuscript.

Regarding the details of POSTRE usage, it was run specifying the “Endocrine” phenotype category and with the rest of default parameters. A potential limitation of the default running mode is that it only considers as disease relevant those genes already associated with the specified phenotypical category (in this case endocrine-related diseases) in established databases, such as OMIM. In this regard, higher sensitivity analyses allowing broader gene inclusion were also explored by disabling the patient phenotype filter (i.e., Gene-PatientPheno option set to No).

All duplications were analysed in POSTRE through the Single SV and Multiple SV Submission interfaces. Regarding the results provided by each mode, the Single SV analysis report included: details of the predicted enhancer adoption events, redefined TAD boundaries (neo-TADs), and additional information for the impacted target genes (illustrated in Figure 2). With respect to the Multiple SV analysis, after uploading all the SVs information through a single txt file (Supplementary Table 1), a set of tables with the aggregated results of the predictions were obtained. POSTRE also allows users to upload their own TAD map to perform the interpretation of the SVs analyzed but this feature was not used, since the TAD map already selected was valid for the locus of interest.

## Results

Thirty three duplications involving *GPR101* (Figure 1) were analyzed using the POSTRE tool, with the phenotype set to “Endocrine” and the running mode set to “Standard” (Figure 2). This configuration integrates enhancer maps derived from ATAC-seq and H3K27ac ChIP-seq data generated from human anterior pituitary cells, alongside RNA-seq profiles specific to this tissue (see Methods). The analysis also relies on prefrontal cortex-derived TAD maps and disease-gene annotations obtained from different databases, such as OMIM^23^ and the Mouse Genome Database^24^. As we established previously, owing to the overall conservation of TADs across cell types, TAD maps from other human tissues can provide informative data on gene regulatory domains and the prediction of long range pathological mechanisms related to SVs^15^.

**Figure 1.**
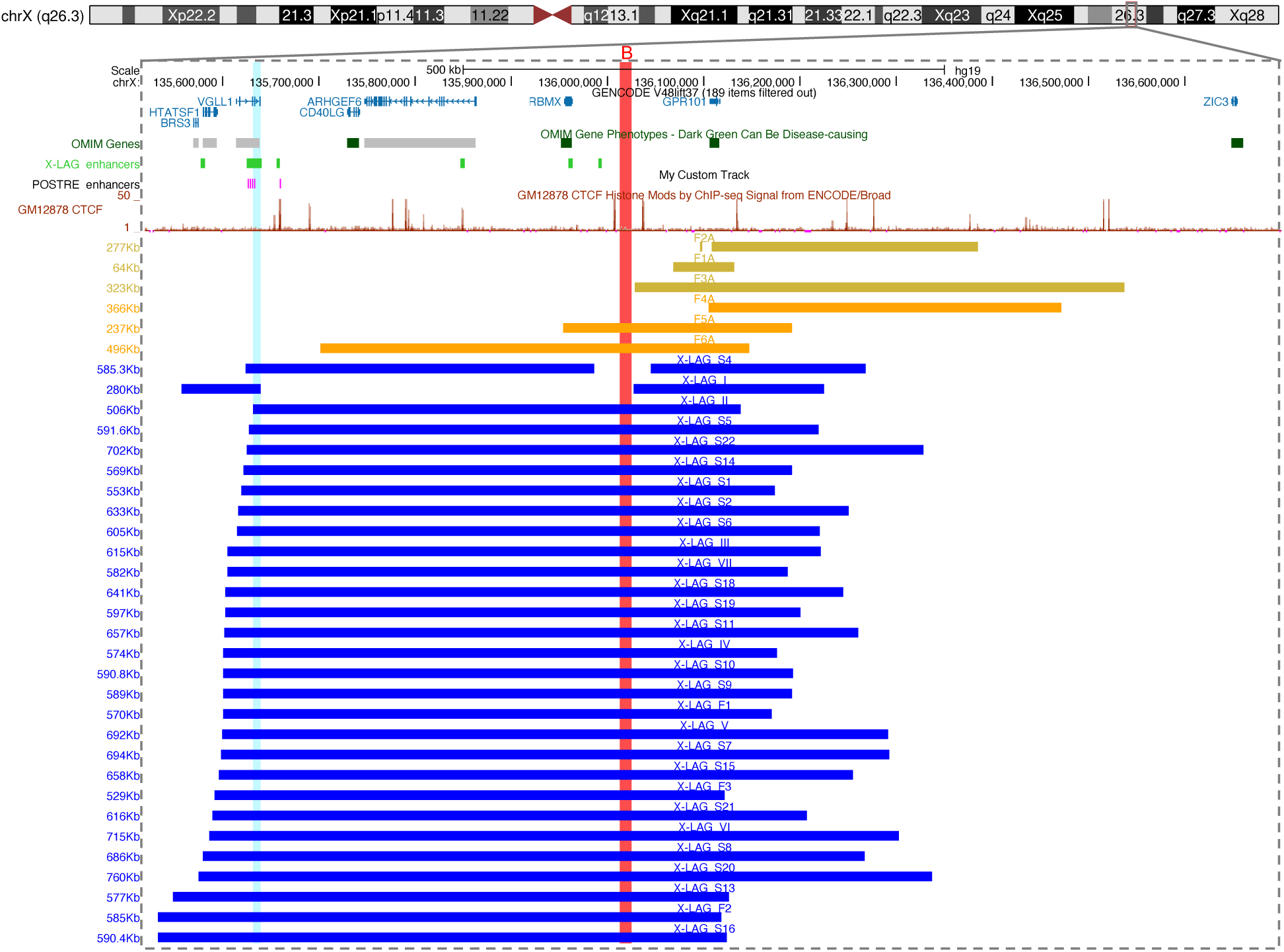
Overview of duplications at the Xq26.3 locus involving *GPR101*, classified by pathogenicity. The genomic region surrounding *GPR101* (chrX:135,500,000-136,700,000, hg19) is shown with annotated protein-coding and OMIM genes (blue and dark green, respectively), CTCF binding sites (orange ChIP-seq track from GM12878 cells), and putative CREs (six we previously described^12^ based on publicly available data^39,40^ given as light green bars and a subset predicted by POSTRE located within or distal to *VGLL1* given as purple bars). The centromeric *GPR101*-TAD boundary is marked by a red vertical bar. Colored bars below represent the extent of individual duplications: yellow for three non-pathogenic duplications reported by Daly et al.^14^ (F1A-F3A), orange for the three newly identified non-pathogenic duplications presented in this study (F4A-F6A), and blue for X-LAG-associated pathogenic duplications, including both continuous and discontinuous (patients S4 and I) rearrangements. Only pathogenic duplications span the *VGLL1* intronic enhancer cluster. The light blue vertical bar highlights the smallest region of overlap of all duplications partially encompassing that CRE. All non-pathogenic duplications, despite partial TAD disruption or inclusion of other CREs such as the *RBMX* enhancer^12^, were predicted by POSTRE to be neutral.

**Figure 2.**
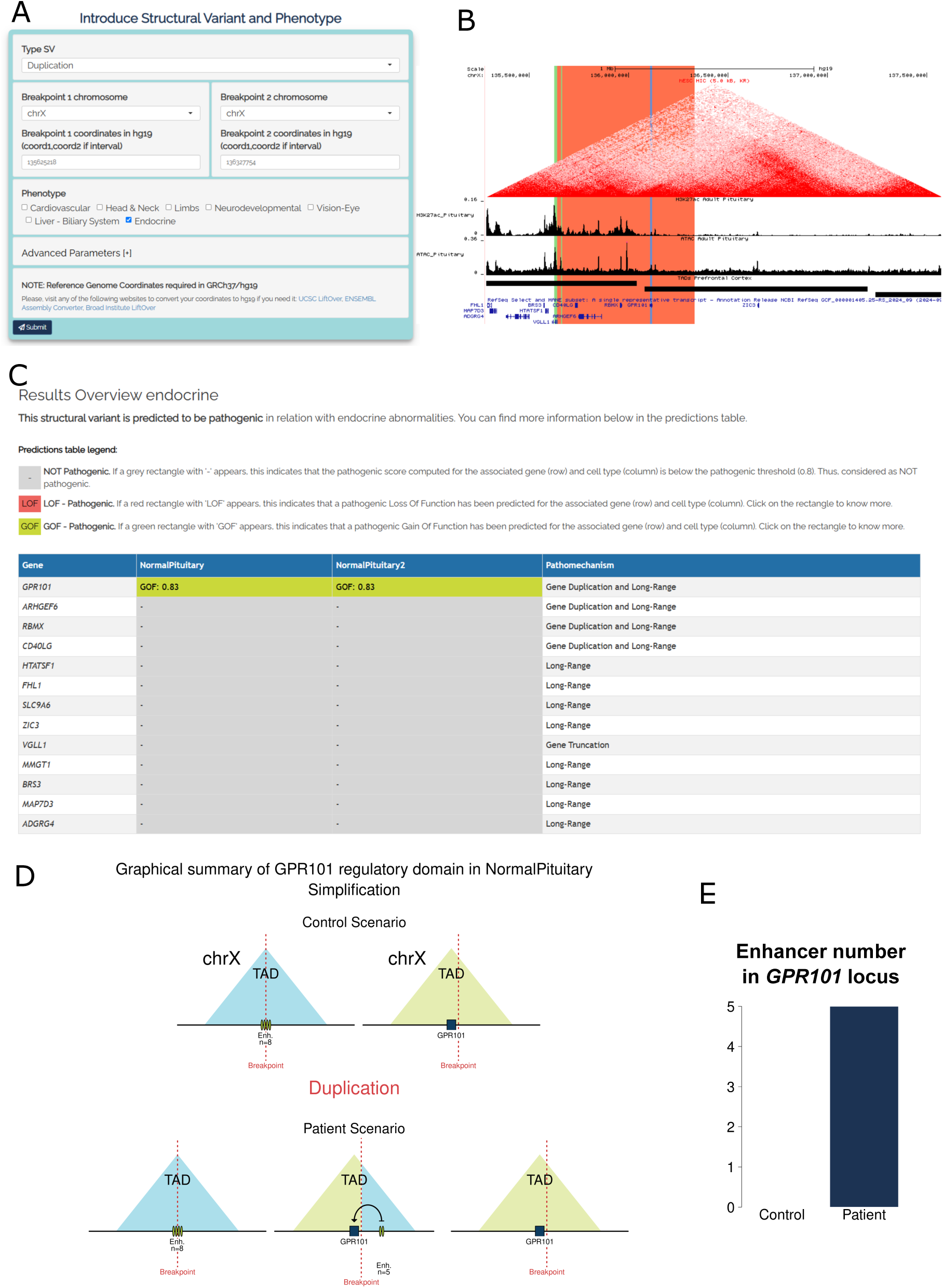
Example of POSTRE output for a pathogenic (X-LAG case S22) duplication at the *GPR101* locus. Panels A-E show part of the results obtained with POSTRE for the analysis of the pathogenic duplication S22. A) Screenshot of POSTRE submission menu for the Single SV analysis mode with the X-LAG case S22 SV coordinates introduced. The duplication was evaluated using POSTRE in Standard mode with the “Endocrine” phenotype selected. This phenotype selection triggers the consideration of anterior pituitary-specific enhancer and expression data. B) Visualization of S22 duplication at the UCSC genome browser. The image was obtained through the adjustment (zoom in) of a UCSC link available from the POSTRE output. The duplicated area is highlighted in red. The session also depicts bigwigs of H3K27ac ChIP-seq and ATAC-seq data from anterior pituitary, TAD maps from the brain prefrontal cortex, and highlights POSTRE predicted active enhancers with green vertical lines. C) Main POSTRE prediction summary table. D) Graphical representation of the SV impact with respect to the *GPR101* regulatory domain. In the control scenario, the existence of eight enhancers can be observed at the TAD containing the *VGLL1*-adjacent enhancers. These eight enhancers include the five *VGLL1*-adjacent enhancers (Figure 4) and three additional ones located at the centromeric side of that TAD (according to the prefrontal cortex TAD map used as reference). However, these latter three enhancers (located >250Kb away from the closest duplication breakpoint) were excluded from downstream considerations as they are never duplicated in any of the reported X-LAG duplications (Figure 1). E) Barplot highlighting the changes in the enhancer number surrounding *GPR101* in the control versus patient alleles. Overall, for S22, POSTRE predicts enhancer adoption and neo-TAD formation involving the *VGLL1*-adjacent enhancers, resulting in a high pathogenicity score and a likely mechanism of *GPR101* misexpression.

In all three of the new non-X-LAG cases, POSTRE predicted the duplications as non-pathogenic (Table 1 and Figure 3). The duplication in individual F4A is entirely contained within the *GPR101* TAD and, accordingly, is not predicted to result in neo-TAD formation—similar to the non-pathogenic duplications we previously reported^14^. By contrast, the duplications in F5A and F6A spanned the centromeric TAD boundary downstream of *GPR101*, resulting in the formation of a neo-TAD. However, unlike in X-LAG associated duplications neither F5A nor F6A included the intronic *VGLL1* enhancer cluster, which is thought to be critical for driving aberrant *GPR101* expression. Both duplications encompassed a putative enhancer located near *RBMX* (eRBMX)^12^, but the POSTRE enhancer calling pipeline does not predict this as one, indicating that this *cis*-regulatory element (CRE) alone might not be sufficient to induce *GPR101* misexpression. As a result, POSTRE predicted these rearrangements to be neutral for *GPR101* regulation despite their inter-TAD configuration. This finding refines the mechanistic interpretation we recently outlined^14^, where the pathogenic importance of TAD boundary disruption was emphasized, but the differential impact of distinct enhancer inclusions was not examined in detail. POSTRE predictions were also benchmarked against the three non-pathogenic *GPR101* duplications that we recently reported (Table 1)^14^. All three were correctly predicted to be non-pathogenic, in line with the original experimental observations using chromatin conformation capture techniques.

**Figure 3.**
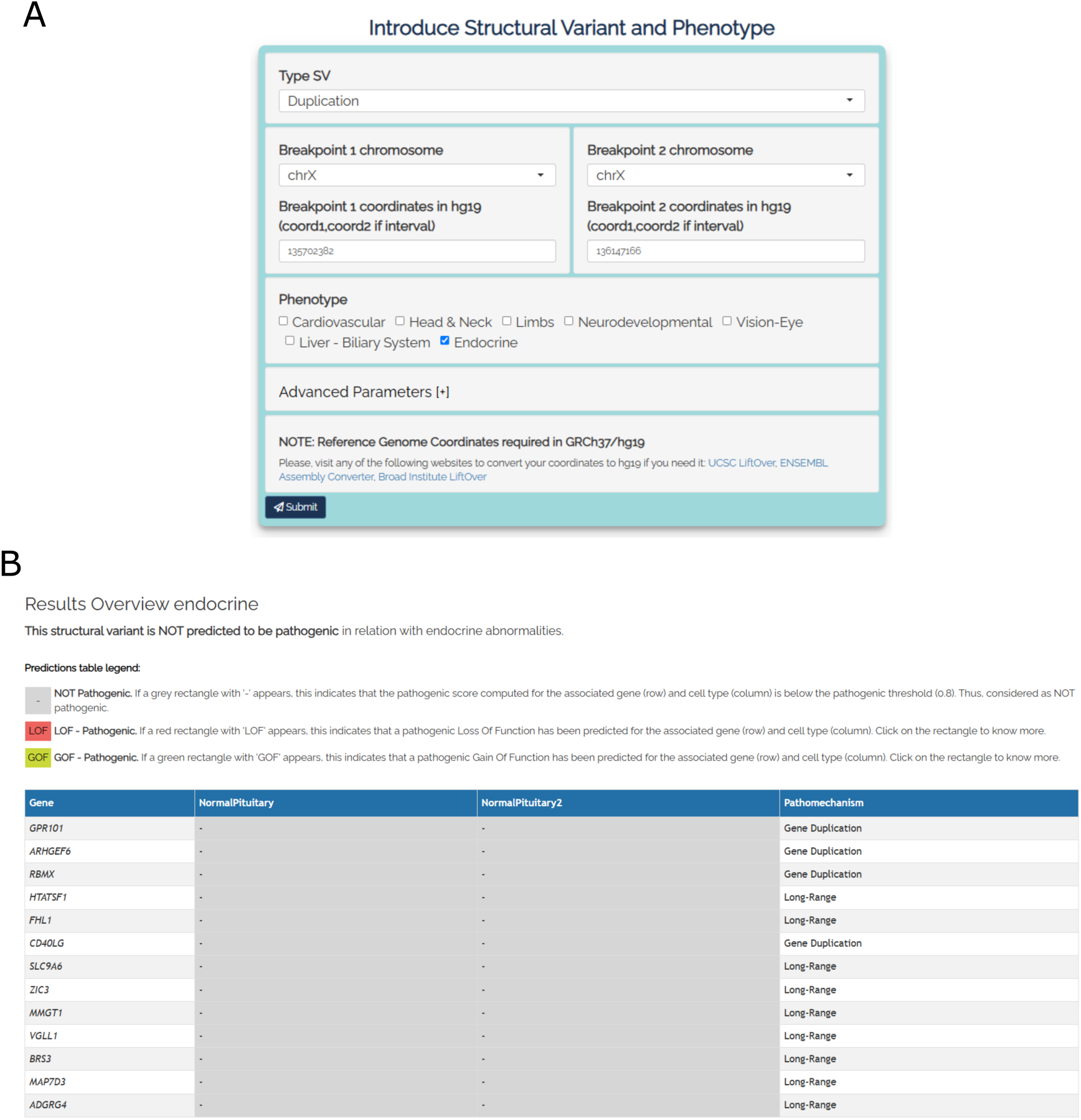
POSTRE output for a non-pathogenic (F6A) duplication at the *GPR101* locus. A) Screenshot of POSTRE submission menu for the Single SV analysis mode with the F6A SV coordinates introduced. The duplication was evaluated using POSTRE in Standard mode with the “Endocrine” phenotype selected. This phenotype selection triggers the consideration of anterior pituitary-specific enhancer and expression data. B) Main POSTRE prediction summary table. It predicts the SV as non-pathogenic.

**Table 1.**
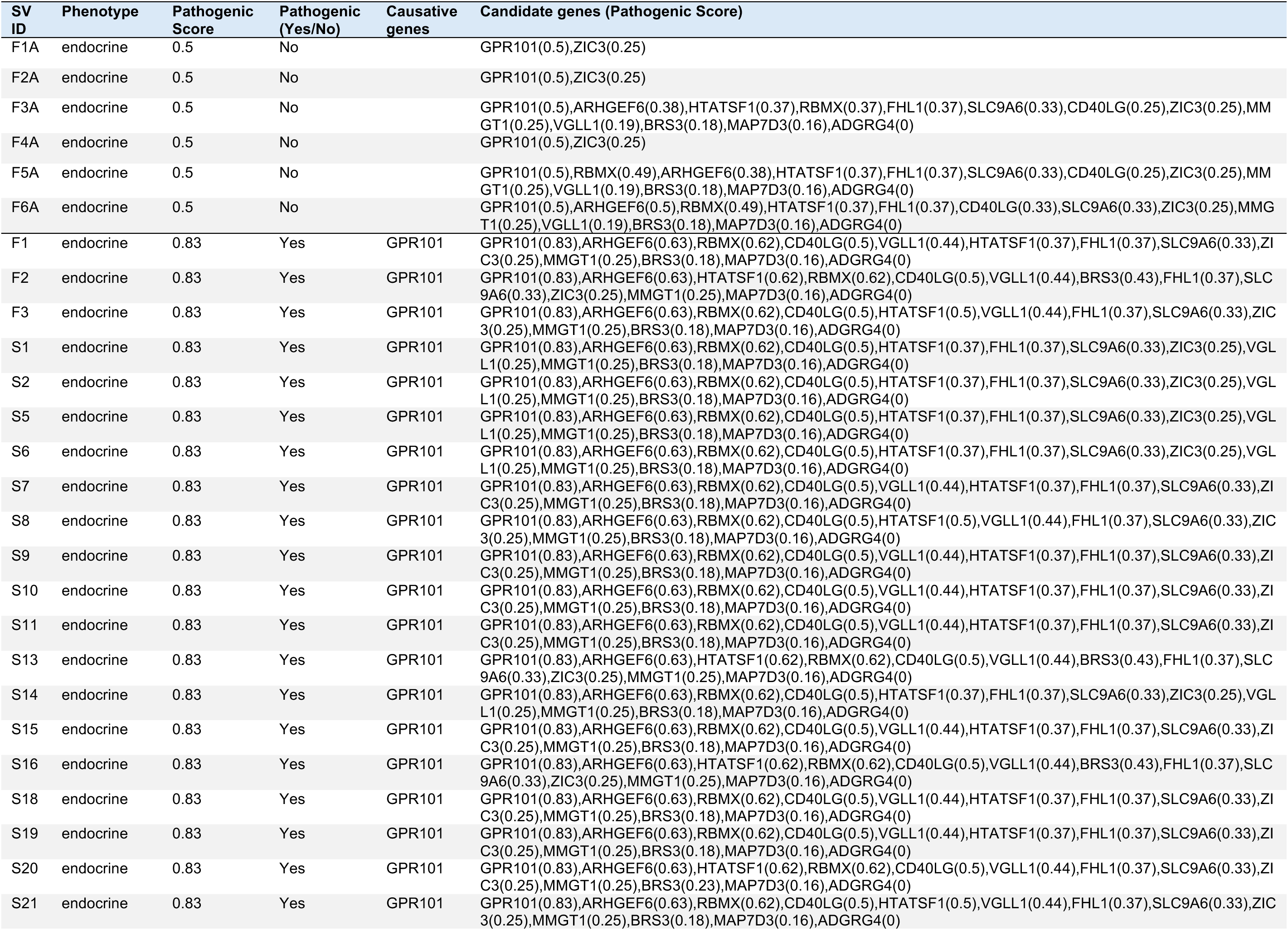
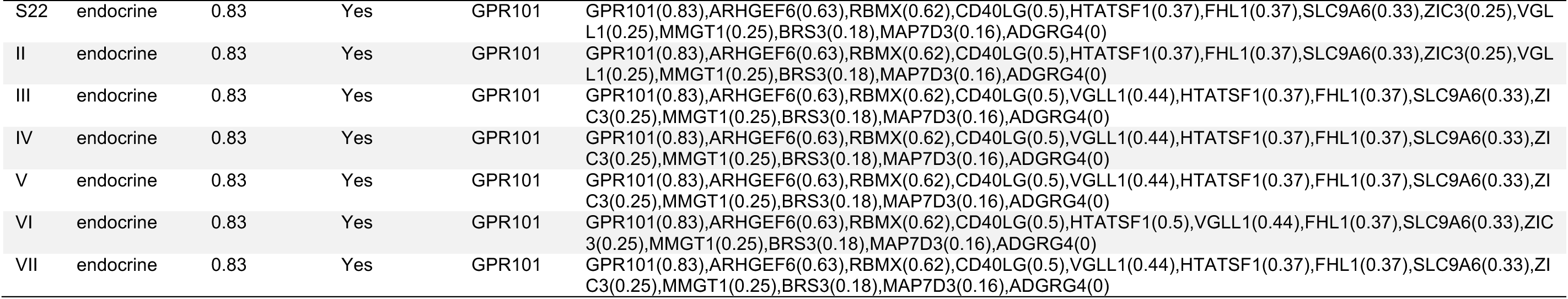
POSTRE output for the Multiple SV analysis. Note: pathogenicity scores for the F3A duplication 515 were also computed 516 by POSTRE for the genes located in the predicted TAD centromeric to *GPR101*. Although the duplication is reported to lie entirely 517 within the *GPR101*-TAD (Figure 1), the TAD map used by POSTRE considers that it encroaches on this centromeric TAD but does 518 not overlap the enhancers located within. This discrepancy reflects a limitation of TAD-calling softwares, as precise annotation of 519 TAD boundaries is often lacking.

| SV ID | Phenotype | Pathogenic Score | Pathogenic (Yes/No) | Causative genes | Candidate genes (Pathogenic Score) |
| --- | --- | --- | --- | --- | --- |
| F1A | endocrine | 0.5 | No |  | GPR101(0.5),ZIC3(0.25) |
| F2A | endocrine | 0.5 | No |  | GPR101(0.5),ZIC3(0.25) |
| F3A | endocrine | 0.5 | No |  | GPR101(0.5),ARHGEF6(0.38),HTATSF1(0.37),RBMX(0.37),FHL1(0.37),SLC9A6(0.33),CD40LG(0.25),ZIC3(0.25),MMGT1(0.25),VGLL1(0.19),BRS3(0.18),MAP7D3(0.16),ADGRG4(0) |
| F4A | endocrine | 0.5 | No |  | GPR101(0.5),ZIC3(0.25) |
| F5A | endocrine | 0.5 | No |  | GPR101(0.5),RBMX(0.49),ARHGEF6(0.38),HTATSF1(0.37),FHL1(0.37),SLC9A6(0.33),CD40LG(0.25),ZIC3(0.25),MMGT1(0.25),VGLL1(0.19),BRS3(0.18),MAP7D3(0.16),ADGRG4(0) |
| F6A | endocrine | 0.5 | No |  | GPR101(0.5),ARHGEF6(0.5),RBMX(0.49),HTATSF1(0.37),FHL1(0.37),CD40LG(0.33),SLC9A6(0.33),ZIC3(0.25),MMGT1(0.25),VGLL1(0.19),BRS3(0.18),MAP7D3(0.16),ADGRG4(0) |
| F1 | endocrine | 0.83 | Yes | GPR101 | GPR101(0.83),ARHGEF6(0.63),RBMX(0.62),CD40LG(0.5),VGLL1(0.44),HTATSF1(0.37),FHL1(0.37),SLC9A6(0.33),ZIC3(0.25),MMGT1(0.25),BRS3(0.18),MAP7D3(0.16),ADGRG4(0) |
| F2 | endocrine | 0.83 | Yes | GPR101 | GPR101(0.83),ARHGEF6(0.63),HTATSF1(0.62),RBMX(0.62),CD40LG(0.5),VGLL1(0.44),BRS3(0.43),FHL1(0.37),SLC9A6(0.33),ZIC3(0.25),MMGT1(0.25),MAP7D3(0.16),ADGRG4(0) |
| F3 | endocrine | 0.83 | Yes | GPR101 | GPR101(0.83),ARHGEF6(0.63),RBMX(0.62),CD40LG(0.5),HTATSF1(0.5),VGLL1(0.44),FHL1(0.37),SLC9A6(0.33),ZIC3(0.25),MMGT1(0.25),BRS3(0.18),MAP7D3(0.16),ADGRG4(0) |
| S1 | endocrine | 0.83 | Yes | GPR101 | GPR101(0.83),ARHGEF6(0.63),RBMX(0.62),CD40LG(0.5),HTATSF1(0.37),FHL1(0.37),SLC9A6(0.33),ZIC3(0.25),VGLL1(0.25),MMGT1(0.25),BRS3(0.18),MAP7D3(0.16),ADGRG4(0) |
| S2 | endocrine | 0.83 | Yes | GPR101 | GPR101(0.83),ARHGEF6(0.63),RBMX(0.62),CD40LG(0.5),HTATSF1(0.37),FHL1(0.37),SLC9A6(0.33),ZIC3(0.25),VGLL1(0.25),MMGT1(0.25),BRS3(0.18),MAP7D3(0.16),ADGRG4(0) |
| S5 | endocrine | 0.83 | Yes | GPR101 | GPR101(0.83),ARHGEF6(0.63),RBMX(0.62),CD40LG(0.5),HTATSF1(0.37),FHL1(0.37),SLC9A6(0.33),ZIC3(0.25),VGLL1(0.25),MMGT1(0.25),BRS3(0.18),MAP7D3(0.16),ADGRG4(0) |
| S6 | endocrine | 0.83 | Yes | GPR101 | GPR101(0.83),ARHGEF6(0.63),RBMX(0.62),CD40LG(0.5),HTATSF1(0.37),FHL1(0.37),SLC9A6(0.33),ZIC3(0.25),VGLL1(0.25),MMGT1(0.25),BRS3(0.18),MAP7D3(0.16),ADGRG4(0) |
| S7 | endocrine | 0.83 | Yes | GPR101 | GPR101(0.83),ARHGEF6(0.63),RBMX(0.62),CD40LG(0.5),VGLL1(0.44),HTATSF1(0.37),FHL1(0.37),SLC9A6(0.33),ZIC3(0.25),MMGT1(0.25),BRS3(0.18),MAP7D3(0.16),ADGRG4(0) |
| S8 | endocrine | 0.83 | Yes | GPR101 | GPR101(0.83),ARHGEF6(0.63),RBMX(0.62),CD40LG(0.5),HTATSF1(0.5),VGLL1(0.44),FHL1(0.37),SLC9A6(0.33),ZIC3(0.25),MMGT1(0.25),BRS3(0.18),MAP7D3(0.16),ADGRG4(0) |
| S9 | endocrine | 0.83 | Yes | GPR101 | GPR101(0.83),ARHGEF6(0.63),RBMX(0.62),CD40LG(0.5),VGLL1(0.44),HTATSF1(0.37),FHL1(0.37),SLC9A6(0.33),ZIC3(0.25),MMGT1(0.25),BRS3(0.18),MAP7D3(0.16),ADGRG4(0) |
| S10 | endocrine | 0.83 | Yes | GPR101 | GPR101(0.83),ARHGEF6(0.63),RBMX(0.62),CD40LG(0.5),VGLL1(0.44),HTATSF1(0.37),FHL1(0.37),SLC9A6(0.33),ZIC3(0.25),MMGT1(0.25),BRS3(0.18),MAP7D3(0.16),ADGRG4(0) |
| S11 | endocrine | 0.83 | Yes | GPR101 | GPR101(0.83),ARHGEF6(0.63),RBMX(0.62),CD40LG(0.5),VGLL1(0.44),HTATSF1(0.37),FHL1(0.37),SLC9A6(0.33),ZIC3(0.25),MMGT1(0.25),BRS3(0.18),MAP7D3(0.16),ADGRG4(0) |
| S13 | endocrine | 0.83 | Yes | GPR101 | GPR101(0.83),ARHGEF6(0.63),HTATSF1(0.62),RBMX(0.62),CD40LG(0.5),VGLL1(0.44),BRS3(0.43),FHL1(0.37),SLC9A6(0.33),ZIC3(0.25),MMGT1(0.25),MAP7D3(0.16),ADGRG4(0) |
| S14 | endocrine | 0.83 | Yes | GPR101 | GPR101(0.83),ARHGEF6(0.63),RBMX(0.62),CD40LG(0.5),HTATSF1(0.37),FHL1(0.37),SLC9A6(0.33),ZIC3(0.25),VGLL1(0.25),MMGT1(0.25),BRS3(0.18),MAP7D3(0.16),ADGRG4(0) |
| S15 | endocrine | 0.83 | Yes | GPR101 | GPR101(0.83),ARHGEF6(0.63),RBMX(0.62),CD40LG(0.5),VGLL1(0.44),HTATSF1(0.37),FHL1(0.37),SLC9A6(0.33),ZIC3(0.25),MMGT1(0.25),BRS3(0.18),MAP7D3(0.16),ADGRG4(0) |
| S16 | endocrine | 0.83 | Yes | GPR101 | GPR101(0.83),ARHGEF6(0.63),HTATSF1(0.62),RBMX(0.62),CD40LG(0.5),VGLL1(0.44),BRS3(0.43),FHL1(0.37),SLC9A6(0.33),ZIC3(0.25),MMGT1(0.25),MAP7D3(0.16),ADGRG4(0) |
| S18 | endocrine | 0.83 | Yes | GPR101 | GPR101(0.83),ARHGEF6(0.63),RBMX(0.62),CD40LG(0.5),VGLL1(0.44),HTATSF1(0.37),FHL1(0.37),SLC9A6(0.33),ZIC3(0.25),MMGT1(0.25),BRS3(0.18),MAP7D3(0.16),ADGRG4(0) |
| S19 | endocrine | 0.83 | Yes | GPR101 | GPR101(0.83),ARHGEF6(0.63),RBMX(0.62),CD40LG(0.5),VGLL1(0.44),HTATSF1(0.37),FHL1(0.37),SLC9A6(0.33),ZIC3(0.25),MMGT1(0.25),BRS3(0.18),MAP7D3(0.16),ADGRG4(0) |
| S20 | endocrine | 0.83 | Yes | GPR101 | GPR101(0.83),ARHGEF6(0.63),HTATSF1(0.62),RBMX(0.62),CD40LG(0.5),VGLL1(0.44),FHL1(0.37),SLC9A6(0.33),ZIC3(0.25),MMGT1(0.25),BRS3(0.23),MAP7D3(0.16),ADGRG4(0) |
| S21 | endocrine | 0.83 | Yes | GPR101 | GPR101(0.83),ARHGEF6(0.63),RBMX(0.62),CD40LG(0.5),HTATSF1(0.5),VGLL1(0.44),FHL1(0.37),SLC9A6(0.33),ZIC3(0.25),MMGT1(0.25),BRS3(0.18),MAP7D3(0.16),ADGRG4(0) |
| S22 | endocrine | 0.83 | Yes | GPR101 | GPR101(0.83),ARHGEF6(0.63),RBMX(0.62),CD40LG(0.5),HTATSF1(0.37),FHL1(0.37),SLC9A6(0.33),ZIC3(0.25),VGLL1(0.25),MMGT1(0.25),BRS3(0.18),MAP7D3(0.16),ADGRG4(0) |
| II | endocrine | 0.83 | Yes | GPR101 | GPR101(0.83),ARHGEF6(0.63),RBMX(0.62),CD40LG(0.5),HTATSF1(0.37),FHL1(0.37),SLC9A6(0.33),ZIC3(0.25),VGLL1(0.25),MMGT1(0.25),BRS3(0.18),MAP7D3(0.16),ADGRG4(0) |
| III | endocrine | 0.83 | Yes | GPR101 | GPR101(0.83),ARHGEF6(0.63),RBMX(0.62),CD40LG(0.5),VGLL1(0.44),HTATSF1(0.37),FHL1(0.37),SLC9A6(0.33),ZIC3(0.25),MMGT1(0.25),BRS3(0.18),MAP7D3(0.16),ADGRG4(0) |
| IV | endocrine | 0.83 | Yes | GPR101 | GPR101(0.83),ARHGEF6(0.63),RBMX(0.62),CD40LG(0.5),VGLL1(0.44),HTATSF1(0.37),FHL1(0.37),SLC9A6(0.33),ZIC3(0.25),MMGT1(0.25),BRS3(0.18),MAP7D3(0.16),ADGRG4(0) |
| V | endocrine | 0.83 | Yes | GPR101 | GPR101(0.83),ARHGEF6(0.63),RBMX(0.62),CD40LG(0.5),VGLL1(0.44),HTATSF1(0.37),FHL1(0.37),SLC9A6(0.33),ZIC3(0.25),MMGT1(0.25),BRS3(0.18),MAP7D3(0.16),ADGRG4(0) |
| VI | endocrine | 0.83 | Yes | GPR101 | GPR101(0.83),ARHGEF6(0.63),RBMX(0.62),CD40LG(0.5),HTATSF1(0.5),VGLL1(0.44),FHL1(0.37),SLC9A6(0.33),ZIC3(0.25),MMGT1(0.25),BRS3(0.18),MAP7D3(0.16),ADGRG4(0) |
| VII | endocrine | 0.83 | Yes | GPR101 | GPR101(0.83),ARHGEF6(0.63),RBMX(0.62),CD40LG(0.5),VGLL1(0.44),HTATSF1(0.37),FHL1(0.37),SLC9A6(0.33),ZIC3(0.25),MMGT1(0.25),BRS3(0.18),MAP7D3(0.16),ADGRG4(0) |

We analyzed 27 X-LAG-associated contiguous duplications at the *GPR101* locus, and POSTRE predicted all 27 to be pathogenic, with each receiving a *GPR101* score of 0.83 (Table 1 and Figure 2). Interestingly, one X-LAG case (patient II) had been previously described as having some unique clinical features^12,17^. Typically, X-LAG is associated with pituitary gigantism due to GH hypersecretion from anterior pituitary adenoma/hyperplasia. Patient II had early-onset GH hypersecretion and overgrowth, but despite many years of detailed clinical follow-up never developed a pituitary tumor. When compared with the other X-LAG cases, the duplication in patient II is also unique. As shown in Figure 4, based on the ATAC peaks enriched in H3K27ac in pituitary cells, POSTRE predicts five enhancers (e1-e5) in the vicinity of *VGLL1*: four in the VGLL1-intronic enhancer (located in introns 2, 3, and 4 of the gene) and one in a more distal telomeric peak (Figure 4 A). These two enhancer loci were previously identified using publicly available, pituitary-specific, human and mouse H3K27ac ChIP-seq and ATAC-seq datasets, respectively^12^. In all the analyzed X-LAG cases, except for patient II and patient S5, all five of the *VGLL1* enhancers were duplicated. The duplication in patient II only involves e4 and e5, which are also the weakest enhancers based on H3K27ac levels (Figure 4B). Regarding S5, which has no atypical features of X-LAG, the duplication includes enhancers e2-e5, some of which are strong by H3K27ac levels, and only excludes the most centromeric enhancer, e1 (Figure 4). These data further emphasize the key importance of the *VGLL1* intronic enhancer cluster and for the first time suggests that the X-LAG phenotype is modulated according to the cumulative strength of these ectopic enhancers acting on the *GPR101* neo-TAD. In the case of patient II, the duplication might lead to a milder enhancer adoption mechanism and, thereby explain the modified clinical phenotype^17,25^.

**Figure 4.**
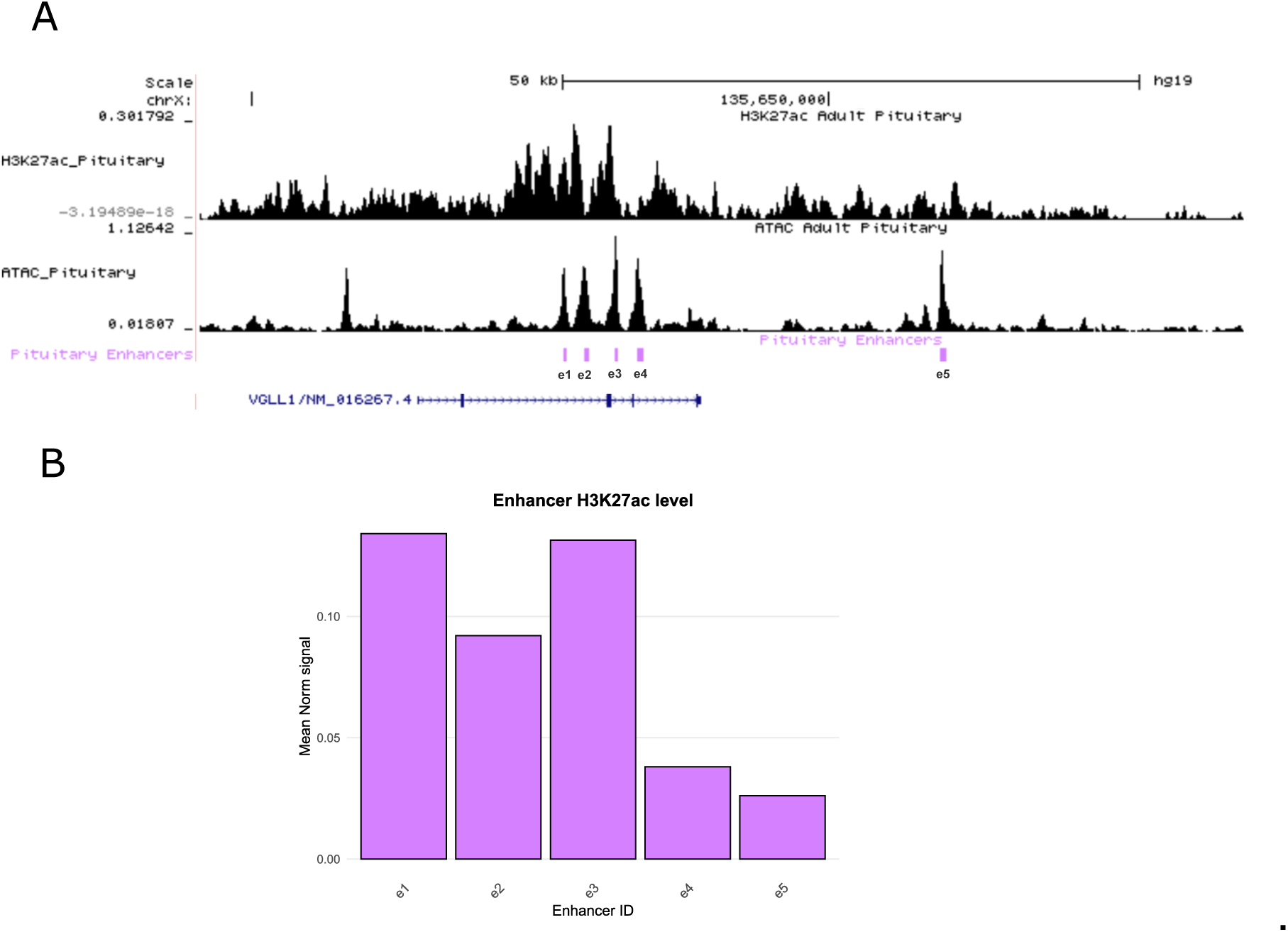
*VGLL1*-adjacent enhancers overview. A) Overview of the *VGLL1*-adjacent enhancers. According to the POSTRE enhancer calling pipeline, five enhancers, that we named here e1-e5, are predicted. B) Barplot of the mean H3K27ac levels (calculated from H3K27ac ChIP-Seq bigwigs) at e1-e5 enhancers. Higher H3K27ac levels are indicators of stronger enhancer activity^41^.

## Discussion

Structural variants altering gene-enhancer communication through TAD disruption are implicated in an emerging group of genomic disorders^3,5–7^. These disorders, which alter regulatory landscapes, present a significant and growing diagnostic challenge in clinical genomics. Their investigation often requires advanced techniques like 4C-seq and HiC, which are not yet optimized for routine diagnostic workflows. Furthermore, integrating these data with other omics datasets (e.g., ChIP-Seq, RNA-Seq) demands specialized genomic and bioinformatics expertise for processing and analysis. This complexity ultimately limits diagnostic yields^26^. In the case of X-LAG, duplications near the *GPR101* locus are known to cause pathogenic rewiring of enhancer-promoter interactions^11,12^. However, as shown previously and expanded on in this study, not all *GPR101*-spanning duplications result in disease, and data on the frequency of 3D genome rearrangements in the general population is unavailable^14^. In parallel, the use of genomic techniques for prenatal screening leads to the identification of SVs of uncertain significance. As not all *GPR101*-related duplications lead to X-LAG, this creates a challenge for clinical geneticists when counselling patients, and misclassification can lead to anxiety and potential misdiagnosis. The ability to reliably distinguish benign from pathogenic duplications at the *GPR101* locus is, therefore, highly clinically relevant, especially in prenatal and pediatric contexts. Our data provide further support for the concept that pathogenicity is not solely determined by TAD disruption or duplication size, but critically depends on inclusion of specific regulatory elements, particularly the pituitary-active enhancer cluster located in the vicinity of the *VGLL1* gene^12^.

In this study, we demonstrate the utility of POSTRE^15^, an *in silico* tool designed to model the regulatory consequences of SVs, at the *GPR101* locus. POSTRE is built upon the integration of several experimentally derived datasets, including ATAC-seq, H3K27ac ChIP-seq, and RNA-seq. These datasets, when combined with available TAD maps, allow the creation of gene-enhancer maps in disease relevant tissues and the possibility of predicting the consequences of their alteration through SVs. While chromatin conformation capture techniques such as 4C-seq and HiC provide direct evidence of three-dimensional genome organization, they are resource-intensive and impractical in routine diagnostic workflows, particularly those that are time-sensitive like prenatal testing. POSTRE complements these experimental approaches by enabling rapid, reproducible predictions of pathogenic enhancer adoption requiring only, from the user, the SV coordinates. When equipped with anterior pituitary-specific datasets, POSTRE correctly classified all known pathogenic X-LAG and non-pathogenic *GPR101* duplications. Crucially, all six duplications—including three from this study and three from our recent work^14^—that were not associated with pituitary dysfunction were classified as benign. This predictive accuracy, coupled with ease of use, highlights its potential for robust diagnostic modeling, particularly when functional assays may be unfeasible or unavailable.

The clinical implications of these findings are especially relevant in time-sensitive contexts such as prenatal diagnosis. In multiple individuals described here and previously, chromosome Xq26.3 duplications were detected through prenatal microarray testing, and were flagged for review based on their proximity to *GPR101* and its association with X-LAG. However, absence of the *VGLL1*-adjacent enhancers, as confirmed by POSTRE modeling, provides strong evidence against a pathogenic interpretation. This not only could aid genetic counseling but also could help to avoid unnecessary monitoring or interventions. In this context, tools like POSTRE provide an objective framework to move toward mechanistically informed variant classification of SVs.

The new pituitary-specific multi-omic dataset used in this study also uncovered important details regarding the *VGLL1*-adjacent enhancers that we identified previously^12^. According to POSTRE enhancer calling criteria, the enhancer cluster identified within *VGLL1* introns is predicted to be formed by four independent enhancers, given the existence of four independent ATAC-seq peaks enriched in H3K27ac. Further research will be needed to assess their independent contributions to *GPR101* expression and their cooperation mode (e.g., additive, synergistic, hierarchical or redundant)^27,28^. In addition, this cluster of *VGLL1* intronic enhancers is accompanied by a single distal enhancer (Figure 4). This information supports the crucial importance of duplication of these enhancers for the generation of the X-LAG phenotype. Since early studies on the genetics of X-LAG, the *VGLL1* locus, along with *GPR101*, formed the smallest regions of overlap (SROs) that were shared by all affected individuals^17,20^ (Figure 1). Since then, we have also identified and tested a series of putative enhancers including one at *RBMX* (eRBMX)^12^. Using POSTRE, the current study indicates the centrality of *VGLL1-*adjacent enhancer elements in the pathogenicity of duplications in X-LAG patients. Interestingly, a partial duplication of a subset of enhancers in patient II led to an atypical form of X-LAG, with severe excess GH hypersecretion in the absence of pituitary tumor formation. No prolactin hypersecretion occurred, which is rare in X-LAG^10,17,29^, and GHRH hypersecretion, that is thought to be responsible for pituitary hyperproliferation in many X-LAG cases^30^, was absent. This case shares many features with the pituitary somatotrope-specific transgenic mouse model that we described in Abboud et al.^31^. That model had gigantism driven by pituitary GH and IGF1 excess, that was driven by an up to 30-fold increase in pituitary *Gpr101* expression (in contrast, *GPR101* is increased 1000s-fold in the tumors of X-LAG patients)^12,32^. Importantly, the *Gpr101* transgenic mouse had a normal pituitary morphology and no evidence of increased proliferation, hyperplasia or tumorigenesis. Taken together, partial duplication of the intronic *VGLL1* enhancer cluster could lead to an incomplete form of X-LAG, hypothetically due to modest, somatotrope-specific elevations in *GPR101* expression. Definitive proof will require further characterization of the enhancer sequences and their functional interactions among themselves (i.e. enhancer cooperation mode), as well as their actions on the *GPR101* promoter. This information would help to further refine the pathogenicity classification of SVs in this region by tools like POSTRE. These observations demonstrate that POSTRE, when equipped with cell-type specific epigenomic and transcriptomic data, can effectively discriminate between pathogenic and benign SVs, even among inter-TAD duplications. Moreover, they reinforce the functional significance of the *VGLL1*-adjacent enhancers in X-LAG and argue against a pathogenic role for the eRBMX CRE, despite its duplication and transcriptional enhancing activity shown in HEK293 cells^12^.

Despite its clear advantages, POSTRE does have limitations. The resolution of predictions is inherently constrained by the quality and granularity of the enhancer and TAD maps it relies upon. In the current version, TAD boundaries are derived from brain prefrontal cortex data and even accounting for strong conservation of TAD boundaries across tissues, may not fully reflect the three-dimensional chromatin architecture of anterior pituitary cells. Future versions will benefit from ongoing efforts to collect and integrate HiC or Micro-C data from pituitary tissues. Additionally, although enhancer annotations were generated for this study using primary adult human pituitary epigenomic datasets, new enhancer maps derived from genomic data at other developmental stages (bulk/single-cell resolution), and improved artificial intelligence strategies to predict enhancer-promoter interactions, could further enhance sensitivity and specificity.

Beyond X-LAG, this study illustrates POSTRE’s broader potential in evaluating SVs across TADopathies affecting the endocrine system, on top of its already proven capabilities to handle this type of alterations in retinal, limb, craniofacial and neurodevelopmental disorders^15,33,34^. Many such conditions are increasingly recognized as the result of disrupted regulatory domains rather than coding mutations^35–38^. Incorporating tissue-relevant data into prediction algorithms allows for more nuanced interpretation of SVs that may otherwise remain unclassified when using *in silico* predictions tools that are agnostic to the cellular context^15,26^. As databases of chromatin and enhancer landscapes expand, and as tools like POSTRE could eventually be integrated into diagnostic pipelines, this will help the field move closer to precision genomic medicine.

In conclusion, the findings presented here establish POSTRE as an effective, interpretable, and scalable tool for the clinical interpretation of SVs at the *GPR101* locus. Moreover, they reinforce the mechanistic model whereby duplications need to span both the *GPR101* TAD boundary and include the *VGLL1*-adjacent enhancers to drive *GPR101* misexpression in X-LAG.

### Ethics approval and consent to participate

Subjects were recruited under the University of Liège Ethics committee approved protocol B707201420418; under the Bioethics Committee of Nicolaus Copernicus University, Toruń, Poland (NCU Committee of Bioethics KB 61/2021); and under the *Eunice Kennedy Shriver* National Institute of Child Health and Human Development, National Institutes of Health protocol 97-CH-0076 (ClinicalTrials.gov: NCT00001595). The study was approved by the Independent Ethics Committee of the IRCCS Humanitas Research Hospital in Rozzano (Milan, Italy) and conformed to the ethical guidelines of the Declaration of Helsinki (approval no. 642/20). Written informed consent was obtained from all the subjects/guardians.

## Data Availability

All data produced in the present study are available upon reasonable request to the authors

## Availability of data and materials

The datasets generated during this study are available on request.

## Authors’ contributions

GT: Conceptualization, Investigation, Writing - Original Draft, Writing - Review & Editing, Visualization, Supervision, Funding acquisition; VS-G: Methodology, Software, Formal analysis, Data Curation, Visualization, Writing - Review & Editing; AG: Investigation, Writing - Review & Editing; MP: Resources, Writing - Review & Editing; CAS: MP: Resources, Writing - Review & Editing; DM: Resources, Writing - Review & Editing; EK: Writing - Review & Editing; AB: Writing - Review & Editing; AGL: Writing - Review & Editing; PP: Resources, Writing - Review & Editing; AR-I: Resources, Writing - Review & Editing; MF: Conceptualization, Writing - Original Draft, Writing - Review & Editing; AFD: Conceptualization, Investigation, Writing - Original Draft, Writing - Review & Editing, Supervision, Funding acquisition.

## Acknowledgements

The authors would like to thank the patients and families involved for their interest, generosity and patience.

## References

1. Dixon JR, Selvaraj S, Yue F, et al. Topological domains in mammalian genomes identified by analysis of chromatin interactions. Nature. Apr 11 2012;485(7398):376-80. doi:10.1038/nature11082

2. Nora EP, Dekker J, Heard E. Segmental folding of chromosomes: a basis for structural and regulatory chromosomal neighborhoods? Bioessays. Sep 2013;35(9):818–28. doi:10.1002/bies.201300040

3. Lupianez DG, Spielmann M, Mundlos S. Breaking TADs: How Alterations of Chromatin Domains Result in Disease. Trends Genet. Apr 2016;32(4):225–237. doi:10.1016/j.tig.2016.01.003

4. Weischenfeldt J, Ibrahim DM. When 3D genome changes cause disease: the impact of structural variations in congenital disease and cancer. Curr Opin Genet Dev. Jun 2023;80:102048. doi:10.1016/j.gde.2023.102048

5. Spielmann M, Lupianez DG, Mundlos S. Structural variation in the 3D genome. Nat Rev Genet. Jul 2018;19(7):453–467. doi:10.1038/s41576-018-0007-0

6. D’Haene E, Vergult S. Interpreting the impact of noncoding structural variation in neurodevelopmental disorders. Genet Med. Jan 2021;23(1):34–46. doi:10.1038/s41436-020-00974-1

7. Valton AL, Dekker J. TAD disruption as oncogenic driver. Curr Opin Genet Dev. Feb 2016;36:34–40. doi:10.1016/j.gde.2016.03.008

8. Matharu N, Ahituv N. Minor Loops in Major Folds: Enhancer-Promoter Looping, Chromatin Restructuring, and Their Association with Transcriptional Regulation and Disease. PLoS Genet. Dec 2015;11(12):e1005640. doi:10.1371/journal.pgen.1005640

9. Rajderkar S, Barozzi I, Zhu Y, et al. Topologically associating domain boundaries are required for normal genome function. Commun Biol. Apr 20 2023;6(1):435. doi:10.1038/s42003-023-04819-w

10. Trivellin G, Daly AF, Faucz FR, et al. Gigantism and acromegaly due to Xq26 microduplications and GPR101 mutation. N Engl J Med. Dec 18 2014;371(25):2363–74. doi:10.1056/NEJMoa1408028

11. Caruso M, Mazzatenta D, Asioli S, et al. Case report: Management of pediatric gigantism caused by the TADopathy, X-linked acrogigantism. Front Endocrinol (Lausanne). 2024;15:1345363. doi:10.3389/fendo.2024.1345363

12. Franke M, Daly AF, Palmeira L, et al. Duplications disrupt chromatin architecture and rewire GPR101-enhancer communication in X-linked acrogigantism. Am J Hum Genet. Apr 7 2022;109(4):553–570. doi:10.1016/j.ajhg.2022.02.002

13. Dimartino P, Zadorozhna M, Yumiceba V, et al. Structural Variants at the LMNB1 Locus: Deciphering Pathomechanisms in Autosomal Dominant Adult-Onset Demyelinating Leukodystrophy. Ann Neurol. Nov 2024;96(5):855–870. doi:10.1002/ana.27038

14. Daly AF, Dunnington LA, Rodriguez-Buritica DF, et al. Chromatin conformation capture in the clinic: 4C-seq/HiC distinguishes pathogenic from neutral duplications at the GPR101 locus. Genome Med. Sep 13 2024;16(1):112. doi:10.1186/s13073-024-01378-5

15. Sanchez-Gaya V, Rada-Iglesias A. POSTRE: a tool to predict the pathological effects of human structural variants. Nucleic Acids Res. May 22 2023;51(9):e54. doi:10.1093/nar/gkad225

16. Beckers A, Lodish MB, Trivellin G, et al. X-linked acrogigantism syndrome: clinical profile and therapeutic responses. Endocr Relat Cancer. Jun 2015;22(3):353–67. doi:10.1530/ERC-15-0038

17. Iacovazzo D, Caswell R, Bunce B, et al. Germline or somatic GPR101 duplication leads to X-linked acrogigantism: a clinico-pathological and genetic study. Acta Neuropathol Commun. Jun 1 2016;4(1):56. doi:10.1186/s40478-016-0328-1

18. Naves LA, Daly AF, Dias LA, et al. Aggressive tumor growth and clinical evolution in a patient with X-linked acro-gigantism syndrome. Endocrine. Feb 2016;51(2):236–44. doi:10.1007/s12020-015-0804-6

19. Trarbach EB, Trivellin G, Grande IPP, et al. Genetics, clinical features and outcomes of non-syndromic pituitary gigantism: experience of a single center from Sao Paulo, Brazil. Pituitary. Apr 2021;24(2):252–261. doi:10.1007/s11102-020-01105-4

20. Trivellin G, Hernandez-Ramirez LC, Swan J, Stratakis CA. An orphan G-protein-coupled receptor causes human gigantism and/or acromegaly: Molecular biology and clinical correlations. Best Pract Res Clin Endocrinol Metab. Apr 2018;32(2):125–140. doi:10.1016/j.beem.2018.02.004

21. Pasinska M, Lazarczyk E, Repczynska A, et al. Clinical Importance of aCGH in Genetic Counselling of Children with Psychomotor Retardation. Appl Clin Genet. 2022;15:27–38. doi:10.2147/TACG.S357136

22. Schmitt AD, Hu M, Jung I, et al. A Compendium of Chromatin Contact Maps Reveals Spatially Active Regions in the Human Genome. Cell Rep. Nov 15 2016;17(8):2042–2059. doi:10.1016/j.celrep.2016.10.061

23. Amberger JS, Bocchini CA, Schiettecatte F, Scott AF, Hamosh A. OMIM.org: Online Mendelian Inheritance in Man (OMIM(R)), an online catalog of human genes and genetic disorders. Nucleic Acids Res. Jan 2015;43(Database issue):D789–98. doi:10.1093/nar/gku1205

24. Bult CJ, Blake JA, Smith CL, Kadin JA, Richardson JE, Mouse Genome Database G. Mouse Genome Database (MGD) 2019. Nucleic Acids Res. Jan 8 2019;47(D1):D801–D806. doi:10.1093/nar/gky1056

25. Burren CP, Williams G, Coxson E, Korbonits M. Effective Long-term Pediatric Pegvisomant Monotherapy to Final Height in X-linked Acrogigantism. JCEM Case Rep. May 2023;1(3):luad028. doi:10.1210/jcemcr/luad028

26. Sanchez-Gaya V, Mariner-Fauli M, Rada-Iglesias A. Rare or Overlooked? Structural Disruption of Regulatory Domains in Human Neurocristopathies. Front Genet. 2020;11:688. doi:10.3389/fgene.2020.00688

27. Blayney JW, Francis H, Rampasekova A, et al. Super-enhancers include classical enhancers and facilitators to fully activate gene expression. Cell. Dec 21 2023;186(26):5826–5839 e18. doi:10.1016/j.cell.2023.11.030

28. Peng Y, Zhang Y. Enhancer and super-enhancer: Positive regulators in gene transcription. Animal Model Exp Med. Sep 2018;1(3):169–179. doi:10.1002/ame2.12032

29. Daly AF, Beckers A. The Genetic Pathophysiology and Clinical Management of the TADopathy, X-Linked Acrogigantism. Endocr Rev. Sep 12 2024;45(5):737–754. doi:10.1210/endrev/bnae014

30. Daly AF, Lysy PA, Desfilles C, et al. GHRH excess and blockade in X-LAG syndrome. Endocr Relat Cancer. Mar 2016;23(3):161–70. doi:10.1530/ERC-15-0478

31. Abboud D, Daly AF, Dupuis N, et al. GPR101 drives growth hormone hypersecretion and gigantism in mice via constitutive activation of Gs and Gq/11. Nat Commun. Sep 21 2020;11(1):4752. doi:10.1038/s41467-020-18500-x

32. Trivellin G, Bjelobaba I, Daly AF, et al. Characterization of GPR101 transcript structure and expression patterns. J Mol Endocrinol. Aug 2016;57(2):97–111. doi:10.1530/JME-16-0045

33. Plaisancie J, Chesneau B, Fares-Taie L, et al. Structural Variant Disrupting the Expression of the Remote FOXC1 Gene in a Patient with Syndromic Complex Microphthalmia. Int J Mol Sci. Feb 25 2024;25(5)doi:10.3390/ijms25052669

34. Hamerlinck L, D’haene E, Van Loon N, et al. Non-coding structural variants identify a commonly affected regulatory region steering *FOXG1* transcription in early neurodevelopment. medRxiv. 2025:2025.03.10.25323301. doi:10.1101/2025.03.10.25323301

35. Carballo-Pacoret P, Carracedo A, Rodriguez-Fontenla C. Unraveling the three-dimensional (3D) genome architecture in Neurodevelopmental Disorders (NDDs). Neurogenetics. Oct 2024;25(4):293–305. doi:10.1007/s10048-024-00774-8

36. Zhang L, Xu M, Zhang W, et al. Three-dimensional genome landscape comprehensively reveals patterns of spatial gene regulation in papillary and anaplastic thyroid cancers: a study using representative cell lines for each cancer type. Cell Mol Biol Lett. Jan 6 2023;28(1):1. doi:10.1186/s11658-022-00409-6

37. Lima AC, Okhovat M, Stendahl AM, et al. Deletion of an evolutionarily conserved TAD boundary impacts spermatogenesis in mice. Biol Reprod. Apr 13 2025;112(4):767–779. doi:10.1093/biolre/ioaf017

38. de Bruijn SE, Fiorentino A, Ottaviani D, et al. Structural Variants Create New Topological-Associated Domains and Ectopic Retinal Enhancer-Gene Contact in Dominant Retinitis Pigmentosa. Am J Hum Genet. Nov 5 2020;107(5):802–814. doi:10.1016/j.ajhg.2020.09.002

39. Mayran A, Khetchoumian K, Hariri F, et al. Pioneer factor Pax7 deploys a stable enhancer repertoire for specification of cell fate. Nat Genet. Feb 2018;50(2):259–269. doi:10.1038/s41588-017-0035-2

40. Vermunt MW, Reinink P, Korving J, et al. Large-scale identification of coregulated enhancer networks in the adult human brain. Cell Rep. Oct 23 2014;9(2):767–79. doi:10.1016/j.celrep.2014.09.023

41. Fulco CP, Nasser J, Jones TR, et al. Activity-by-contact model of enhancer-promoter regulation from thousands of CRISPR perturbations. Nat Genet. Dec 2019;51(12):1664–1669. doi:10.1038/s41588-019-0538-0

