## Supplementary Figure 1 for "Distinguishing benign from pathogenic duplications involving *GPR101* and *VGLL1*-adjacent enhancers in the clinical setting with the bioinformatic tool POSTRE"

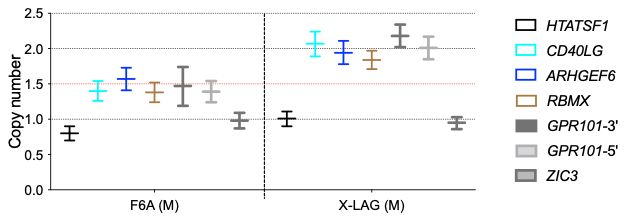

**Supplementary Figure 1. Results of copy number (CN) ddPCR assays in subject F6A.**

This male individual harbors a duplication that includes *GPR101* and its neighboring centromeric genes, up to *CD40LG*. As control, a typical X-LAG subject (shown here is a familial male case^10^) was analyzed in the same experiment. Blood-derived DNA was used for analysis in all subjects. The calculated CN along with Poisson-based 95% confidence intervals (CN range bars) are shown for each gene. Note that two assays located at the 5′ and 3′ end of *GPR101* coding sequence were employed. The dotted red lines crossing the y axis at CN value 1.5 represents the threshold for duplication in males.
